# Locus Coeruleus Integrity in Older Adults with and without Chronic Pain

**DOI:** 10.1101/2021.11.02.21265820

**Authors:** Tyler R. Bell, Carol E. Franz, Lisa T. Eyler, Christine Fennema-Notestine, Olivia K. Puckett, Stephen M. Dorros, Matthew S. Panizzon, Rahul C. Pearce, Donald J. Hagler, Michael J. Lyons, Asad Beck, Jeremy A. Elman, William S. Kremen

**Author notes:** Dr. Elman and Dr. Kremen are joint senior authors.

## Abstract

The locus coeruleus (LC) is a brainstem region involved in regulating pain. Chronic pain is common in older adulthood, but no studies have examined its association with the LC in humans. We used neuromelanin-sensitive imaging to study differences in LC integrity in older adults with and without chronic pain. Chronic pain was assessed in community-dwelling men from the Vietnam Era Twin Study of Aging (VETSA) in 3 study waves covering an average of 12 years. Pain was self-reported on the SF-36 Bodily Pain Scale. Chronic pain was defined as moderate to severe pain severity at the current and at least one prior wave; 17% had chronic pain (*n*=80). At the third wave, 481 participants (mean age=67.57) underwent neuromelanin-sensitive MRI scans from which we calculated an LC contrast-to-noise ratio (LC_CNR_) – an index of LC integrity. We examined associations between chronic pain and LC_CNR_ (in the rostral LC and caudal regions) with generalized estimating equations after adjusting for age, race, education, depressive symptoms, medical comorbidities, and opioid medication use. Individuals with chronic pain had .35 standard deviation lower rostral LC_CNR_ (95% CI: -.62 to -.05) compared to those without chronic pain. No differences in the caudal LC_CNR_ were detected. Chronic pain was associated with decreased rostral LC integrity in older adults. Differences in the rostral LC, rather than caudal LC, suggest the association between lower LC integrity and chronic pain may be related to pain processing in cortical regions where rostral LC projections typically connect.

## Introduction

Approximately 27%-33% of older adults experience chronic pain, i.e., pain lasting three or more months [16]. Such pain is linked to increased risk of cognitive impairment [23,54], negative affective symptoms [2,12,27], sleep disruption [15], and reduced physical function in older adults [47]. Identifying brain regions related to chronic pain in older adults may help uncover reasons for poor pain modulation and other clinical symptoms. Human neuroimaging studies have found evidence of reduced gray matter volume and white matter integrity [2;10]. Animal studies suggest that the brainstem might also play an important role in pain [33]. However, this region has not been examined in pain studies of humans. A brainstem region of particular interest is the locus coeruleus (LC), a small nucleus (30-60mm^3^) that serves as the central projection site for norepinephrine (NE) [49].

The LC is essential to acute pain processing. After tissue damage, nociceptive signals are gated (allowed to ascend or not) at inter-neuronal spaces throughout the dorsal horn and further modulated by caudal LC projections to pre- and post-synaptic terminals [37]. Nociceptive signals received by the caudal LC then modulate sensory and affective pain processing in cortical regions. Sensory pain processing involves caudal and rostral LC-NE projections to the thalamus and parietal cortices that determine whether one feels pain and where. Affective pain processing involves rostral LC-NE projections to the prefrontal cortex, amygdala, and insula [38] that determine how one feels and responds to pain. Because of the essential roles of the LC in acute pain processing, chronic pain may involve poorer structural integrity of this region [43].

Different theories of chronic pain and neurodegeneration may suggest how chronic pain and LC integrity may be associated. *Active theory* suggests that pain initiates signaling from the spinal cord to the brain that in turn results in neuroinflammatory processes and brain atrophy, due to increased cytokine levels [26,31,52] and/or increased tau and beta-amyloid formation [13,17,24,25]. *Passive theory* posits that many age-associated neurodegenerative diseases harm pain-processing brain regions, a consequence of which is the experience of chronic pain. Chronic pain following major neurodegenerative diseases such as dementia, Parkinson’s disease, and motor neuron disease are in support of this theory [18]. Lastly, *developmental theory* suggests that early brain characteristics put people at risk for chronic pain due to poor pain modulation, as shown in prospective studies [5,50].

Here we examined the association between chronic pain and LC integrity in older adults for the first time. LC structural integrity was based on an MRI measure of neuromelanin signal [33]. We hypothesized that people with chronic pain would show lower LC integrity compared to people without chronic pain. We examined the rostral and caudal LC regions separately, given the importance of their respective connections to cortical sensory/affective pain processing and nociceptive gating in the dorsal horn [42]. Neuromelanin signals from the middle LC were combined with the rostral signals as both regions have overlapping projection sites and similar changes related to age and neurodegenerative disease [9,10,37].

## Methods

### Participants

Participants were from the Vietnam Era Twin Study of Aging (VETSA), a longitudinal study of male twins assessed at average ages 56, 62, and 68 – an approximately 12-year time span [34]. All participants served in the military at some time during the Vietnam era (1965-1975). The majority did not serve in Southeast Asia, and about 80% reported no combat exposure. This sample is representative of general community-dwelling men in their age range on demographics, health, and other lifestyle factors based on Centers for Disease Control and Prevention data [44]. Previous studies have described the methodology and available data in the VETSA project [34,35]. Data are publicly accessible to researchers through data authorization (http://www.vetsatwins.org/for-researchers/). Procedures of the VETSA were approved by the Institutional Review Boards at the University of California, San Diego, and Boston University.

LC imaging was introduced at wave 3 (2016 to 2019). Of eligible MRI participants, 485 (92.4%) met standard MRI inclusion criteria (e.g., no metal in the body) and completed imaging of the LC. Four were excluded due to poor image quality. The final sample consisted of 481 individuals with an average age of 67.52 years (*SD* = 2.60, range = 61.96 to 71.72). Demographic characteristics of this sample can be seen in Table 1.

**Table 1.** Demographics of sample (*n* = 481).

| | % | $n$ | $M$ | $SD$ | Range |
| --- | --- | --- | --- | --- | --- |
| Age |  |  | 67.52 | 2.60 | 61.96 to 71.72 |
| Chronic pain | 16.6% | 80 |  |  |  |
| Race |  |  |  |  |  |
| Non-Hispanic White | 88.1% | 498 |  |  |  |
| Education (years) |  |  | 13.98 | 2.07 | 8 to 20 |
| Medical Comorbidities |  |  |  |  |  |
| 0 | 17.5% | 84 |  |  |  |
| 1 | 33.1% | 159 |  |  |  |
| 2+ | 49.5% | 238 |  |  |  |
| Depressive Symptoms |  |  | 6.84 | 6.90 | 0 to 47.00 |
| Opioid Medications |  |  | 1.36 | 0.88 | 0.14 to 9.00 |
| 0 | 95.0% | 457 |  |  |  |
| 1+ | 5.0% | 24 |  |  |  |

Notes. Chronic pain was defined as pain on the SF-36 Bodily Pain Intensity item (Question 22) greater than mild ( $>3/6$ ) in two or more study waves. Medical comorbidities included major health conditions from the Charlson Index. Depressive symptoms were measured from the Center for Epidemiological Studies Depression scale. Opioid medications were coded as the number of opioid medications recorded during the medical history interview.

### Measures

#### Chronic pain

Pain was assessed using the SF-36 Quality of Life (Version 1.0) Bodily Pain Scale [53], which was given at each assessment wave. Specifically, we used the pain severity item (Question 22) asking participants, “How much pain severity have you had during the past 4 weeks”. Severity was rated on a 6-point Likert-type scale from “None” (1),“Very Mild” (2),“Mild” (3),“Moderate” (4),“Severe” (5),“Very Severe” (6). Chronic pain was defined as moderate to very severe pain (values 4-6 on the scale) at wave 3 and the previous participating wave. Pain reported over two or more waves (approximately 6 to 12 years) fits common definitions of chronic pain as moderate to severe pain severity [32] that lasts 3 or more months [16]. People with no pain or pain during only one wave were classified as not having chronic pain.

#### LC MRI Acquisition and Processing

We previously described our procedures for MRI imaging of the LC [20]. In brief, participants completed scans in GE 3T Discovery 750x scanners (GE Healthcare, Waukesha, WI, USA) with an eight-channel phased-array head coil. Oblique axial FSE-T1-weighted images were then obtained (TR = 600 ms; TE = 14 ms; flip angle = 90°; matrix = 512 × 320; FOV = 220 mm; pixel size 0.42 × 0.68 mm; 10 slices; slice thickness = 2.5 mm; interslice gap = 1 mm).

Each image was manually marked by two of four experienced raters according to methods described in detail previously [22]. Briefly, signal intensities were derived from manually marked regions of interest (ROIs) on three axially oriented slices corresponding to rostral, middle, and caudal LC. On each slice, a 3mm^2^-voxel crosshair-shaped ROI was placed over left and right LC and a 10 mm^2^ square reference ROI was placed in the pontine tegmentum (PT). Mean signal was then extracted from each ROI. The values from left and right LC were highly correlated (*r*s > .98) and were subsequently averaged for each slice. Finally, LC contrast-to-noise ratio (LC_CNR_) values were calculated for each slice as LC_CNR_=(LC_intensity_ -PT_intensity_)/PT_intensity_. Higher LC_CNR_ values are thought to reflect better LC structural integrity [33]. For each individual, we calculated the rostral and caudal LC_CNR_: we averaged LC_CNR_ values from the rostral and middle slices to derive the final rostral LC_CNR_ value and used the LC_CNR_ value from the caudal marked slice to derive the final caudal LC_CNR_ value.

#### Covariates

Variables that could confound an association between chronic pain and LC_CNR_ were included as covariates. These included age, race/ethnicity (non-Hispanic white versus other), education (years), depressive symptoms indexed using the 20-item Center of Epidemiological Studies scale (CES-D) [40], medical comorbidities (total number of reported major health conditions from a modified Charlson Index [14]), and the use of opioid medication based on reports of medications taken in the medical history interview (yes/no).

### Statistical Analysis

Descriptive statistics are shown in Table 1, and we compared key covariates between people with and without chronic pain to identify potential confounding variables. General estimating equations (GEEs) using unstructured correlation matrices were conducted using SAS (version 9.4) to examine the association between chronic pain and LC_CNR_, providing effect estimates while accounting for the correlation of observations within twin pairs. Separate models were conducted for rostral and caudal LC_CNR_ with each outcome regressed onto chronic pain status and adjusted for covariates. Statistical significance was set to an *α* of .05, and 95% confidence intervals were calculated. Models were conducted using bootstrapping with 1000 resamples to provide robust estimates.

## Results

### Descriptive Statistics

In participants completing LC_CNR_ measurement, 16.6% (*n* = 80) reported chronic pain. Compared with individuals who did not report chronic pain, participants reporting chronic pain had more depressive symptoms (*t*(97.44) = 4.30, *p* <.001, *d* = .87), more medical comorbidities (*t*(98.26) = 5.16, *p* = .013, *d* = 1.04), and were more likely to be using opioid medications (*χ*^2^(1) = 25.67, *p* <.001, *η*^2^ = .23; 16.3% versus 2.8%), but did not differ in age (*p* = .069).

### Main Results

Shown in Table 2 and Figure 1, participants with chronic pain had significantly lower rostral LC_CNR_ (*b* = -.35, 95% CI: -.62 to -.05, *β* = -.35, *p* = .042) than those without chronic pain. Of the covariates, older age was associated with lower rostral LC_CNR_ (*b* = -.04, 95% CI: -.05 to - .01, *β* = -.13, *p* = .027). Race/ethnicity, education, depressive symptoms, medical comorbidities, and opioid use were not significantly associated with rostral LC_CNR_ (*p*s > .05).

**Table 2.** Main results of general estimating equations predicting rostral LC_CN_ with bootstrapping.

| Variable | $b$ | 95%CI | $p$ | $\beta$ |
| --- | --- | --- | --- | --- |
| Rostral $LC_{CNR}$ (z-score) | | | | |
| Chronic pain | -.35 | -.62 to -.05 | .042 | -.13 |
| Age | -.04 | -.05 to -.01 | .027 | -.14 |
| non-Hispanic White | .05 | -.28 to .38 | .778 | .01 |
| Education (years) | .03 | -.01 to .08 | .150 | .07 |
| Medical Comorbidities | .03 | -.06 to .09 | .497 | -.04 |
| Depressive Symptoms | -.01 | -.02 to .01 | .483 | -.05 |
| Opioid Medication Use | .45 | .07 to .98 | .061 | .14 |

Notes. CNR = contrast to noise; LC = locus coeruleus. The rostral $LC_{CNR}$ value was calculated as the average CNR value from the rostral and middle LC regions. The model estimates were conducted using bootstrapping with 1000 resamples. Chronic pain was defined as pain on the SF-36 Bodily Pain Intensity item (Question 22) greater than mild ( $>3/6$ ) in two or more study waves. Medical comorbidities included major health conditions from the Charlson Index. Depressive symptoms were measured from the Center for Epidemiological Studies Depression scale. Opioid medications were coded as the number of opioid medications recorded during the medical history interview.

**Figure 1.**
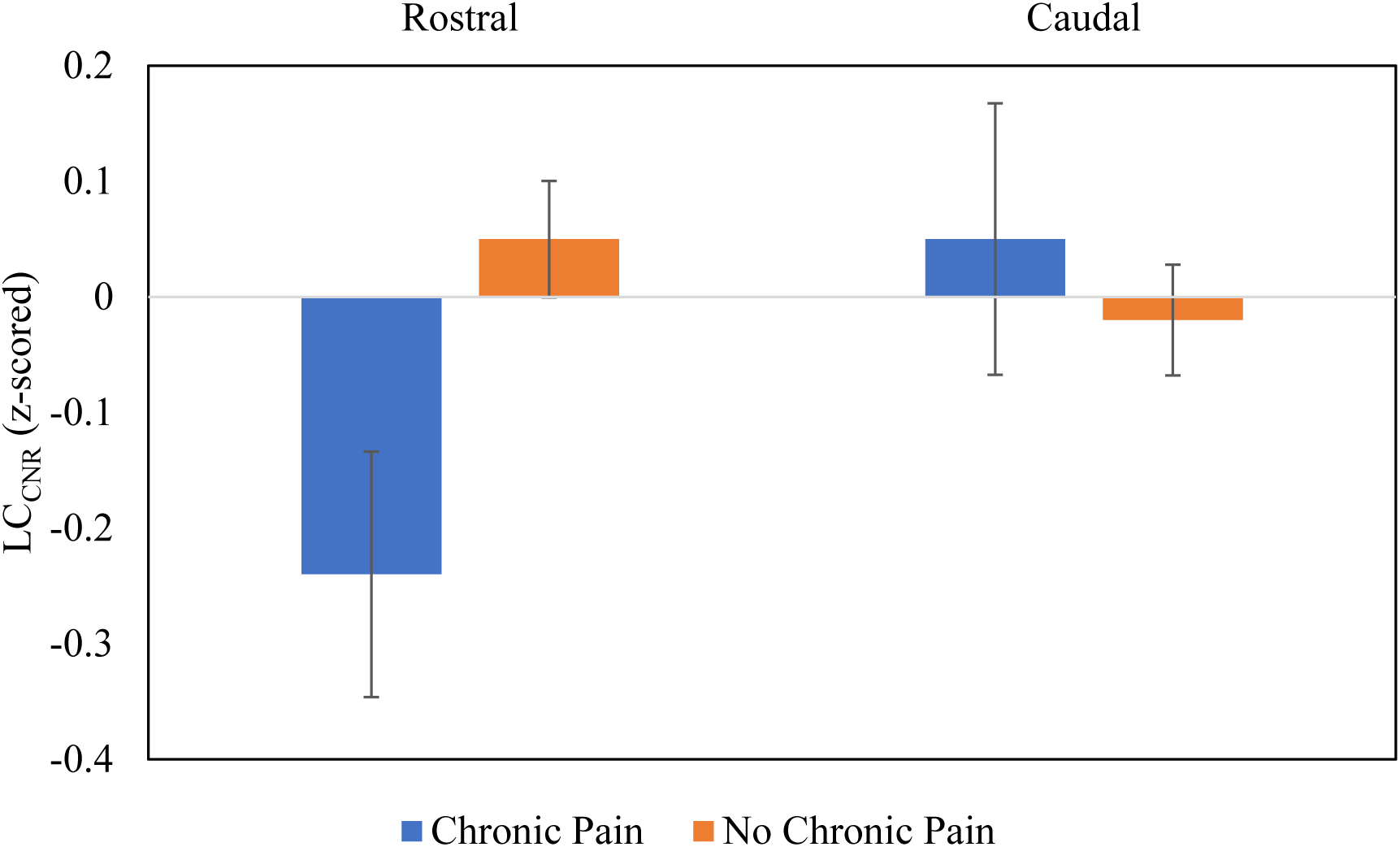
Differences in the rostral and caudal locus coeruleus integrity measured by locus coeruleus contrast to noise ratio (LC_CNR_) in people with and without chronic pain. On this scale, 0 represents the average LC_CNR_ for the whole sample, units away from 0 represent standard deviations from the sample average. Rostral LC_CNR_ was the average CNR from the rostral and middle LC regions.

As shown in Table 3 and Figure 1, participants with chronic pain did not differ from participants without chronic pain on caudal LC_CNR_ (*p* = .755). No covariate was significantly associated with caudal LC_CNR_ (*p*s > .05).

**Table 3.** Main results of general estimating equations predicting caudal LC_CN_ with bootstrapping.

| Variable | *b* | 95%CI | *p* | $\beta$ |
| --- | --- | --- | --- | --- |
| Caudal LC<sub>CNR</sub> (z-score) |  |  |  |  |
| Chronic pain | -.05 | -.20 to .40 | .755 | -.02 |
| Age | -.02 | -.05 to .01 | .065 | -.12 |
| non-Hispanic White | -.13 | -.45 to .18 | .409 | -.04 |
| Education (years) | .02 | -.02 to .07 | .296 | .05 |
| Medical Comorbidities | .03 | -.05 to .11 | .418 | .05 |
| Depressive Symptoms | -.002 | -.03 to .01 | .848 | -.02 |
| Opioid Medication Use | .16 | -.35 to .72 | .524 | .05 |

Notes. CNR = contrast to noise; LC = locus coeruleus. The model estimates were conducted using bootstrapping with 1000 resamples. Chronic pain was defined as pain on the SF-36 Bodily Pain Intensity item (Question 22) greater than mild (>3/6) in two or more study waves. Medical comorbidities included major health conditions from the Charlson Index. Depressive symptoms were measured from the Center for Epidemiological Studies Depression scale. Opioid medications were coded as the number of opioid medications recorded during the medical history interview.

## Discussion

Our analyses highlighted an important link between chronic pain and region-specific LC integrity in older adults. Overall, individuals with chronic pain had lower structural integrity in the rostral LC compared to those without chronic pain. This difference was not accounted for by age, depressive symptoms, medical comorbidities, or opioid use. No significant associations were observed for the caudal LC region. To our knowledge, this is the first examination of associations between chronic pain and LC integrity in humans.

An association between chronic pain and rostral LC integrity adds to previous literature implicating the LC in human pain modulation. Anatomical studies have linked the LC to several important brain regions involved in sensory and affective pain processing, and the LC has connections to the dorsal horn of the spine where nociceptive signals are gated [37]. Animal studies have shown a causal role of the LC in pain modulation. In rats, for example, stimulation of the LC suppresses pain while lesions to the LC increase pain sensitivity [37]. Other studies found that drugs that target the LC-NE system through gabapentinoids [28] and serotonin-norepinephrine reuptake inhibitors markedly inhibit neuropathic pain in rats with ablated or constricted nerve injury [1]. Such effects can be reversed with the application of NE antagonists [28]. Early evidence consistent with the LC-NE system’s role in human pain comes from successful treatment with similar medications [4,46]. Although our study cannot determine causation, our findings are consistent with evidence that the LC is related to human pain.

Results were region-specific. Caudal LC integrity was not different in people with chronic pain compared to people without chronic pain. As mentioned, the rostral LC has more projection sites to cortical regions involved in the sensory and affective processing of pain, while the caudal LC has more projection sites in the dorsal horn involved in gating of nociceptive signals [37]. Although the previous literature has shown a strong role of dorsal horn activity in chronic pain [31], our study suggests that the rostral region is most related to chronic pain in humans.

Although chronic pain classification was based on longitudinal data at two earlier waves, LC integrity was measured only at the last wave, leaving directionality unresolved. As highlighted by active, passive, and developmental theories of chronic pain and neurodegeneration, multiple temporal directions are possible. The active theory would suggest that lower rostral LC integrity is a consequence of global brain atrophy related to chronic pain and neuroinflammation [42]. The LC may even be particularly vulnerable due to its small size and proximity to the spinal cord, where neuroinflammation is thought to originate [36,52]. Passive theory would suggest that lower rostral LC integrity arises due to age-associated neurodegenerative diseases, resulting in chronic pain. Many brainstem regions are the first affected be neurodegenerative diseases, which may make the LC vulnerable. For example, Alzheimer’s disease is characterized by pathology in the LC at its earliest stages [11]. Lastly, lower rostral LC integrity could represent differences from earlier brain development, making chronic pain more likely as we age [5,50]. It seems likely that associations between LC integrity and chronic pain are bidirectional and accounted for, in part, by elements of each of these theories. However, directionality and causation remain to be resolved with longitudinal assessment and experimental animal modeling.

Our measure of LC integrity may have functional implications to consider, especially if lower integrity reflects structural degeneration. Damaged LC regions produce more tonic levels of NE to projections sites which activate pain-processing brain regions [3,30,51]. In animal models, tonic NE release from the LC has been shown to promotes affective pain processing in the amygdala and anterior cingulate cortex specifically [55]. This has also been shown to result from inducing pain-related injury [48]. Increased tonic LC-NE outputs could explain why people with chronic pain show more tonic pupillary responses [8,39], which correlate with greater pain intensity generally [29]. Although more work will be needed to discern NE patterns, it could be that people with chronic pain have higher tonic discharge relating to lower rostral LC integrity, thereby increasing cortical pain processing. Put another way, the link between chronic pain and rostral, but not caudal, LC integrity suggests more top-down rather than bottom-up regulation of the response to pain.

Our finding that older adults with persistent pain have lower rostral LC integrity might explain other pain-related symptoms. In particular, the LC is a key structure involved in arousal and modulation of cognitive function. For example, our previous work in this sample showed that people with lower rostral LC integrity had greater daytime sleep-related dysfunction [21] – a symptom frequently reported in people with chronic pain [19]. Furthermore, people with chronic pain have worse cognitive performance than those without chronic pain [6,7]. Our prior work has shown better rostral LC integrity may be related to better cognitive performance and reduced risk of amnestic mild cognitive impairment [21]. In other studies, lower LC integrity has also been associated with major depressive disorder [45], a condition more common in people with chronic pain. As such, LC integrity might play a role in other phenomena associated with chronic pain, although the causal direction of the LC-pain association or the associations of chronic pain with other LC-related phenomena remains unclear.

Our study has some limitations and strengths. First, as mentioned, we cannot address causality, although evidence thus far suggests that the relationship may be bidirectional. Also, we cannot ascertain the pathology contributing to lower LC integrity in this population without histological data, although inflammatory processes are suspected. Secondly, our sample is entirely male and primarily non-Hispanic white, making generalizability to women and racial/ethnic minorities uncertain. Regarding strengths, we used a novel MRI approach to provide one of the first in vivo analyses of chronic pain and LC integrity while adjusting for several possible confounding factors.

Chronic pain is a costly condition in later life, reducing quality of life, cognitive function, and physical abilities. Identifying the biological mechanisms behind its inception and continuation is thus a worthwhile goal and could identify potential treatment targets to promote healthier aging trajectories. Such work is of increasing importance as the number of adults over age 65 will triple by 2050, along with cases of chronic pain [41]. Our study adds to previous findings linking the LC and pain, showing that older adults with chronic pain have lower rostral LC integrity than their counterparts without chronic pain. Many questions remain, but more data using neuromelanin MRI imaging and further animal modeling will be crucial, especially for determining the role of exact pathways and LC-NE discharge patterns.

## Data Availability

VETSA data can be accessed through data requests submitted online (http://www.vetsatwins.org/for-researchers/). Request for additional information regarding the specific use of data and data analytic methods may be sent to the first author.

## Author contributions

Description of authors’ roles: All authors contributed to the conception or design of the work; or the acquisition, analysis, or interpretation of the work. All authors helped to revise the work for intellectual content and approved the final draft. All authors are accountable for all aspects of the work ensuring accuracy and integrity.

## Sponsors’ role

This work was supported by the National Institute on Aging at the National Institutes of Health grant numbers R01s AG050595, AG022381, AG037985, AG062483, and P01 AG055367, and K01AG063805.

## Acknowledgments

The content is the responsibility of the authors and does not necessarily represent official views of the NIA, NIH, or VA. The U.S. Department of Veterans Affairs, Department of Defense; National Personnel Records Center, National Archives and Records Administration; National Opinion Research Center; National Research Council, National Academy of Sciences; and the Institute for Survey Research, Temple University provided invaluable assistance in the creation of the VET Registry. The Cooperative Studies Program of the U.S. Department of Veterans Affairs provided financial support for development and maintenance of the Vietnam Era Twin Registry. We would also like to acknowledge the continued cooperation and participation of the members of the VET Registry and their families.

## Conflict of Interest Declaration

The authors declare the absence of known competing financial or personal relationships that could have influenced the work reported in this paper.

## Notes

### Competing Interest Statement

The authors have declared no competing interest.

### Author Declarations

The Institutional Review Board of the University of California San Diego approved the procedures of this work.

